# Impact of azithromycin mass drug administration on the antibiotic-resistant gut microbiome: a randomized, controlled trial

**DOI:** 10.1101/2021.03.02.20215632

**Authors:** Harry Pickering, John D. Hart, Sarah Burr, Richard Stabler, Ken Maleta, Khumbo Kalua, Robin L. Bailey, Martin J. Holland

## Abstract

**Background:** Mass drug administration (MDA) with azithromycin is the primary strategy for global trachoma control efforts. Numerous studies have reported secondary effects of MDA with azithromycin, including reductions in childhood mortality, diarrhoeal disease and malaria. Most recently, the MORDOR clinical trial demonstrated that MDA led to an overall reduction in all-cause childhood mortality in targeted communities. There is however concern about the potential of increased antimicrobial resistance in treated communities.

**Methods:** This study evaluated the impact of azithromycin MDA on the prevalence of gastrointestinal carriage of macrolide-resistant bacteria in communities within the MORDOR Malawi study, additionally profiling changes in the gut microbiome after treatment. For faecal metagenomics, 60 children were sampled prior to treatment and 122 children after four rounds of MDA, half receiving azithromycin and half placebo.

**Findings:** The proportion of bacteria carrying macrolide resistance increased after azithromycin treatment; the effect was enhanced in children treated within six months of sampling. Diversity and global community structure of the gut was minimally impacted by treatment, however abundance of several species was altered by treatment. Notably, the putative human enteropathogen *Escherichia albertii* was more abundant after treatment.

**Interpretation:** The impacts of MDA with azithromycin, including increased carriage of macrolide-resistant bacteria, were enhanced in children treated more recently, suggesting effects may be transient. Increased abundance of enteropathogenic *Escherichia* species after treatment requires further, higher resolution investigation. Future studies should focus on the number of treatments and administration schedule to ensure clinical benefits continue to outweigh costs in antimicrobial resistance carriage.

**Funding:** Bill and Melinda Gates Foundation

## Introduction

Mass drug administration (MDA) with azithromycin has been the cornerstone of trachoma control programs since the 1990s and the advent of the SAFE strategy^1^. There has been considerable research since into the secondary effects of community-wide azithromycin distribution. A study from The Gambia was the first to report ancillary benefits^2^. They found all-cause illness, fever, diarrhoea and vomiting were reduced for at least one-month post-treatment compared with topical tetracycline. Similarly, a study from Nepal found reductions in impetigo and diarrhoea up to ten days post-treatment^3^. In 2009, Porco *et al* reported a 50% reduction in all-cause mortality in children aged 1-9 years in Ethiopia in communities given azithromycin MDA^4^. This work has been expanded upon in studies across sub-Saharan Africa, demonstrating decreases in diarrhoea, malaria and all infectious mortality^5–11^.

Alongside these benefits, there has been evidence of negative effects, primarily the emergence and increasing prevalence of antimicrobial resistance. A study of Aboriginal communities in Australia reported short-term reductions in the prevalence of nasopharyngeal *Streptococcus pneumoniae* carriage, but significant increases in macrolide-resistance in identified isolates^12^. Both effects had waned considerably 2-months post-treatment, however prevalence of macrolide-resistant isolates remained elevated above pre-treatment levels at six months follow-up. Further studies have supported this increase in macrolide-resistant nasopharyngeal *S. pneumoniae*^3,13,14^ as well as in faecal *Escherichia coli*^15–17^, with evidence of macrolide and non-macrolide antimicrobial resistance in the latter. There is limited evidence of macrolide resistance in the primary clinical target of azithromycin MDA, *Chlamydia trachomatis*^18–20^, or the unintended target *Plasmodium falciparum*^8^.

Better defining the impact of azithromycin MDA on childhood morbidity and mortality was the aim of the MORDOR (Mortality reduction after oral azithromycin) clinical trial, which randomised 1533 communities across three sub-Saharan African countries to four biannual rounds of azithromycin treatment or placebo^21^. Azithromycin treatment led to an overall reduction in all-cause childhood mortality in targeted communities of 13·5% at 24-month follow-up, although the effect size varied between countries (Malawi; 5·7%, Niger; 18·1%, Tanzania, 3·4%). Secondary analyses found the impact of treatment to be most pronounced in the first three months post-treatment and in children aged 1-5 months^21,22^. Despite Niger reporting the most significant effect on childhood mortality, there were no significant differences in the causes of mortality between the two study arms in this country^23^. In both the treatment and placebo arms, malaria (28%), pneumonia (16%) and diarrhoea (14-15%) accounted for the majority of verbal autopsy-confirmed deaths. All-cause mortality was reduced by 9% after treatment in Malawi, while this decrease was not significant, secondary analysis suggested pneumonia and diarrhea or HIV/AIDS mortality were the drivers of this effect^24^.

Findings from studies nested within MORDOR in Niger supported previous work that reported increases in antimicrobial resistance after azithromycin MDA. The proportion of macrolide-resistant nasopharyngeal *S. pneumoniae* isolated was four times greater after treatment^25^. Additionally, macrolide-resistance determinants were more prevalent in the gut after treatment. Doan *et al* further explored the impact of treatment on antimicrobial resistance and gut microbiome composition in Nigerien communities through metagenomics^26,27^. They found increased prevalence of macrolide resistance after treatment, prevalence was approximately seven times greater after both four and eight bi-annual treatments. Additionally, after eight rounds of treatments, prevalence was also increased for other antimicrobial resistance classes, most prominently β-lactams. Treatment also had a long-term impact on the gut microbiome at 24-month follow-up, reducing diversity, as previously reported, and altering abundance of specific species^28,29^. Notably, prevalence of the rarely identified human pathogen *Campylobacter upsaliensis* decreased after treatment. However, the majority of affected species have little known role in gut health or pathogenicity. In contrast, findings from a study nested within MORDOR in Malawi which profiled the gut microbiome by 16S rRNA sequencing, found no change in diversity after treatment and limited impact on individual genera, with only a minor increase in *Prevotella* reported^30^.

This study evaluated the impact of azithromycin MDA on the prevalence of gastrointestinal carriage of macrolide-resistant bacteria in communities within the MORDOR Malawi study site, where the observed reduction in childhood mortality was considerably less than in Niger. Additionally, this study investigated changes in the gut microbiome after treatment.

## Methods

### Study design and participants

The study design has previously been described^31^. Briefly, the randomization unit for the MORDOR trial in Malawi was defined as the catchment area of a Health Surveillance Assistant (HSA), approximately 1,000 total population. Communities with population <200 or >2000 on a pre-baseline census were excluded. Thirty communities were randomly selected for follow-up as part of studies of child morbidity and antimicrobial resistance (Figure 1). The randomization was stratified to produce 6 communities in each of the 5 geographical zones of Mangochi District for geographical generalisability as well as for logistical reasons. Samples collected in the Makinjira zone were eligible for the current study of antimicrobial resistance determined by whole-genome-sequencing. Biannual census updates were performed, and communities received study drug in the same treatment rounds as the MORDOR mortality trial^21^.

**Figure 1.**
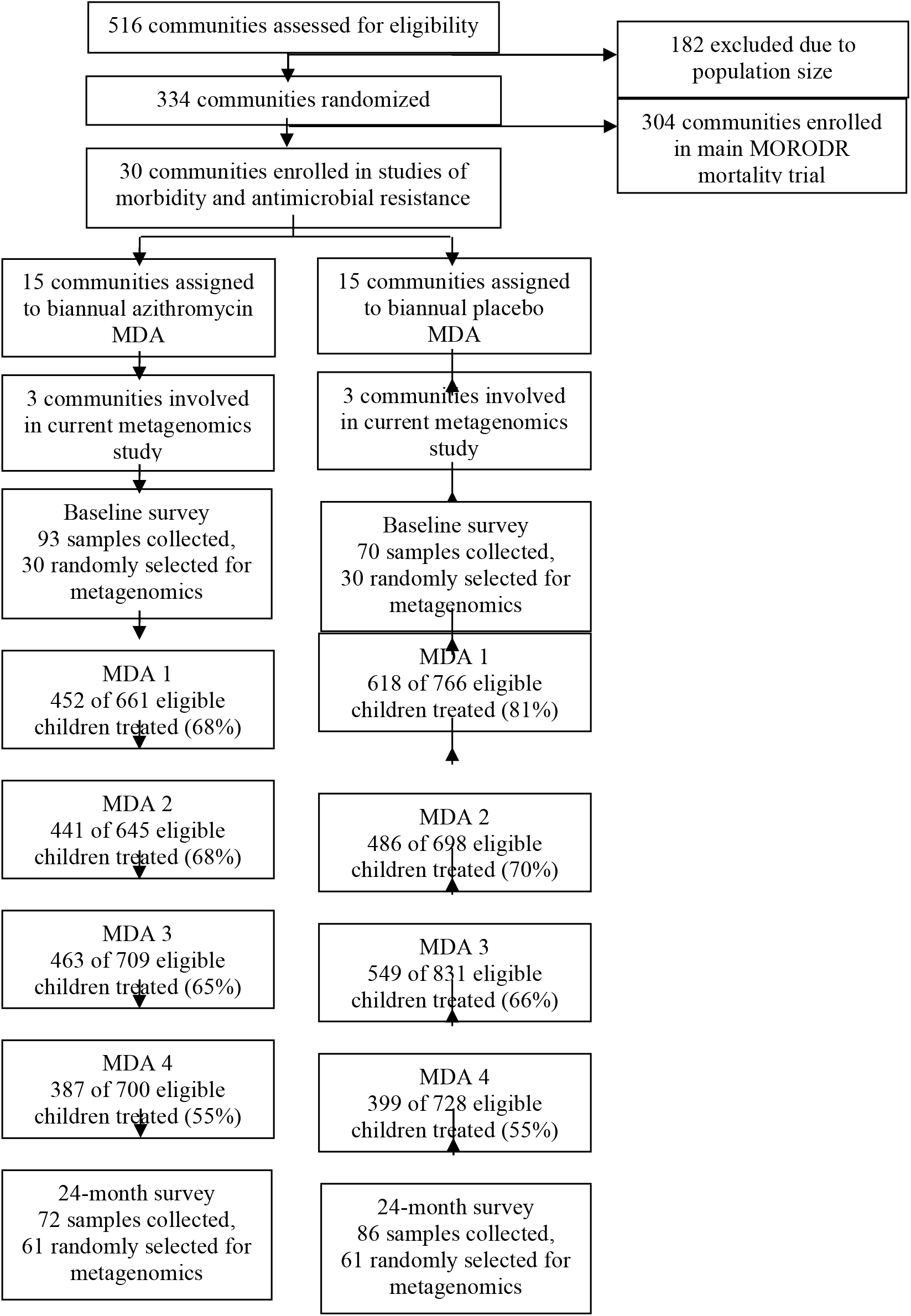
Study design. Flowchart illustrating the study protocol.

Trained local nursing staff explained the procedures and study and at the baseline, 12-month and 24-month follow-up visits, 40 children aged 1-59 months per community were randomly selected for stool sampling.

### Intervention

All children aged 1-59 months and weighing ≥3·8kg were eligible for treatment at each of 4 biannual mass distributions. Azithromycin was administered at a dose of 20mg/kg. Children old enough to stand received an approximate dose estimated from their height and younger children were weighed. Distribution of the drug took place after sample collection was complete and was performed by the HSAs and fieldworkers conducting house-to-house visits. Guardians were asked to inform the HSA of any adverse events that occurred within 7 days of receiving study drug. HSAs subsequently informed the study team.

### Sample collection

Sample collection took place during the baseline visit (May-July 2015) and at 12-month and 24-month visits (April-June 2016 and 2017 respectively), approximately 6 months after the second and fourth treatment rounds (Figure 1). Parents or guardians who provided consent for sample collection were provided with a stool sample collection kit and were given instruction on how to collect the sample. Samples were returned to the field team immediately after collection and were held on wet ice until transport to the lab (within 8 hours of collection). Once at the lab, samples were stored at −80°C until further processing.

### Metagenomic sequencing

Bacterial DNA was extracted from 182 stool samples using the QIAamp PowerFecal DNA Kit (Qiagen) and eluted in EB Buffer. Concentration and purity of the extracts were assessed using a Nanodrop ND8000 (Thermo Scientific) and a subset of samples were processed on a Genomic Screentape using an Agilent 2200 Tapestation to assess integrity of the extracts. One hundred ng extracted DNA was used to generate whole genome libraries using the NEBNext® Ultra™ II FS DNA Library Prep Kit for Illumina (New England Biolabs) alongside NEBNext® Multiplex Oligos for Illumina® (New England Biolabs). The average sizes of the resulting libraries were assessed using D1000 Screentapes on an Agilent 2200 Tapestation and concentrations were assessed using the Qubit™ dsDNA HS Assay Kit. The libraries were normalized to 4nM and pooled into 4 pools of 37 libraries and 1 pool of 38 libraries. The average sizes of the pools were assessed using High Sensitivity D1000 Screentapes on an Agilent 2200 Tapestation and concentrations were assessed using the Qubit™ dsDNA HS Assay Kit. Each pool was subsequently sequenced on a NextSeq 500 using NextSeq® 500/550 High Output Kit v2 (Illumina) in a 150-cycle paired-end configuration.

### Metagenomic analysis

Raw reads were filtered, trimmed and error-corrected using AfterQC^32^. Filtered reads were assembled into contigs using MEGAHIT^33^. Kraken^34^ was used for taxonomic assignment of assembled contigs against complete bacterial, archaeal, and viral genomes from RefSeq^35^, as of November 2017. ABRicate (github.com/tseeman/abricate) was used to screen contigs for antimicrobial resistance genes against the ResFinder database^36^.

### Outcome measures

Pre-specified primary outcome measures included the prevalence of carriage of macrolide resistance in the stool, at 24-month follow-up, as determined by genetic determinants. Secondary outcome measures included microbial diversity in the intestinal microbiome, at 24-month follow-up, as measured using next generation sequencing.

### Statistical analysis

Alpha diversities and principal component values were compared by linear regression, adjusting for age. Species abundance profiles and proportion of bacteria carrying antimicrobial resistance genes were compared by zero-inflated negative binomial regression, adjusting for age. Time since last treatment analyses were further adjusted for number of treatments received. P-values were corrected for multiplicity of testing using the Benjamini-Hochberg procedure^37^.

### Ethical approval

Ethical approval for the MORDOR trials in Malawi was obtained from the College of Medicine Research and Ethics Committee, College of Medicine, University of Malawi (Malawi) and the London School of Hygiene and Tropical Medicine Observational/Interventions Research Ethics Committee, London School of Hygiene and Tropical Medicine (UK). Written, informed consent was obtained from the guardians of participants of all participating children. Illiterate guardians provided a thumb print to acknowledge consent.

## Results

### Participants

Thirty samples per treatment arm at baseline and 61 samples per treatment arm at 24-month follow-up were randomly selected for metagenomic sequencing (Figure 1). Age and sex were comparable between treatment arms (Table 1). As expected, gut microbial diversity, based on Shannon’s H, increased with age (p = 0·237×10-9). Microbial composition, based on principal component analysis (PCA) of Bray-Curtis dissimilarity, changed significantly with age (Principal component (PC)1; p = 0·331×10-11, PC2; p = 0·003). Sex did not significantly impact gut microbial diversity (p = 0·754) or composition (PC1; p = 0·504, PC2; p = 0·300). Additionally, at baseline, study arms were equivalent in gut microbial diversity (p = 0·596) and composition (PC1; p = 0·472, PC2; p = 0·894). All analyses were adjusted for age of participants and multiplicity of testing.

**Table 1.** Patient demographics.

| Variable | Baseline |  | 24-month follow-up |  |
| --- | --- | --- | --- | --- |
|  | Placebo (n = 30) | Azithromycin (n = 30) | Placebo (n = 61) | Azithromycin (n = 61) |
| Median age in years (range) | 2·87 (0·74 – 4·59) | 2·88 (0·81 – 4·91) | 2·45 (0·22 – 5·01) | 2·17 (0·29 – 4·98) |
| Female (%) | 16 (53·33) | 12 (40·00) | 27 (44·26) | 30 (49·18) |

### Antimicrobial resistance profile after treatment

The majority of individuals (180/182) carried at least one bacterium carrying macrolide resistance, with no difference by arm or time (Table 2). Bacteria most frequently carrying macrolide resistance were *Enterococcus* (*E. cecorum* and *E. faecium*), *Bifidobacterium* (*B. breve, B. longum* and *B. kashiwanohense*) and *Streptococcus* (species unknown). No changes in prevalence of carriage of macrolide resistance were found in individual species over time.

**Table 2.**
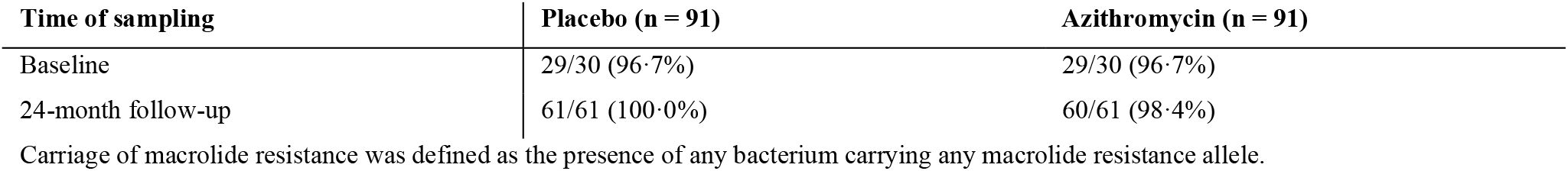
Prevalence of carriage of macrolide resistance bacteria Time of sampling Placebo (n = 91)

| Time of sampling | Placebo (n = 91) | Azithromycin (n = 91) |
| --- | --- | --- |
| Baseline | 29/30 (96·7%) | 29/30 (96·7%) |
| 24-month follow-up | 61/61 (100·0%) | 60/61 (98·4%) |

Carriage of macrolide resistance was defined as the presence of any bacterium carrying any macrolide resistance allele.

Due to the high baseline prevalence of carriage of bacteria with macrolide resistance, we compared the proportion of bacteria carrying macrolide resistance. The proportion of bacteria carrying macrolide resistance increased significantly after treatment (p = 0·827×10-7) (Figure 2a). There were no significant differences in non-macrolide antimicrobial resistance after treatment, although an increase in prevalence of bacteria carrying aminoglycoside (p = 0·064) and trimethoprim (p = 0·059) resistance was close to significance (Figure 2b). Carriage of macrolide resistance was not correlated with either aminoglycoside (Pearson’s r = −0·154) or trimethoprim resistance (r = − 0·153), carriage of resistance to these two non-macrolide antibiotics was correlated (r = 0·849).

**Figure 2.**
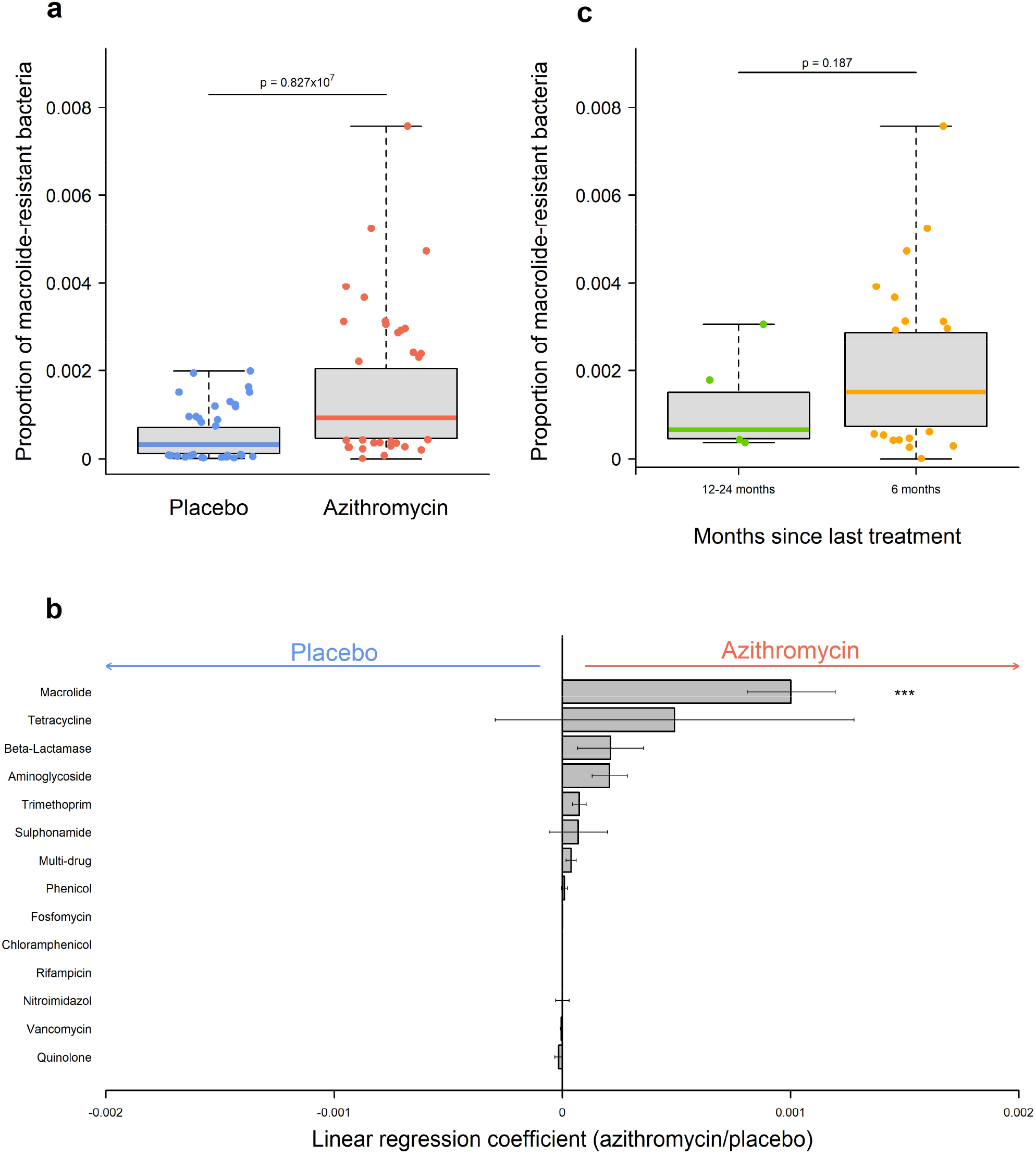
Antimicrobial resistance profile in children receiving either placebo or azithromycin treatment. a. The proportion of macrolide-resistance bacteria at 24-month follow-up in children receiving either placebo (blue) or azithromycin (red). b. Univariate analysis of antibiotic classes with increased evidence of resistance (linear regression coefficient > 0) or decreased evidence of resistance (linear regression coefficient < 0) after azithromycin treatment. c. The proportion of macrolide-resistance bacteria at 24-month follow-up in children receiving azithromycin 6 months or 12-24 months previously. P-values were considered significant at <0.05 and are denominated as follows: *p < 0·05.

Most studies on the impact of azithromycin treatment have focused on changes in the days or weeks post-treatment. We therefore examined the relationship between time since treatment and macrolide resistance carriage. Sixty percent of individuals in the azithromycin arm received treatment 6-months before sample collection at 24-month follow-up (37/61), individuals whose last treatment occurred 12-, 18- or 24-months prior to follow-up sample collection were combined (24/61). All analyses were adjusted for number of treatments received. There was a trend towards increased proportion of bacteria carrying macrolide resistance in individuals treated 6-months prior to sample collection compared to those treated 12-24-months prior, this did not reach significance (p = 0·187) (Figure 2c).

### Changes in the gut microbiome after treatment

To further explore changes in the gastrointestinal flora, we determined the impact of treatment on gut microbiome composition. In the azithromycin arm, gut microbial diversity was unchanged after treatment (p = 0·454) (Figure 3a). Treatment did not impact global gut microbial composition (PC1; p = 0·699, PC2; p = 0·551) (Figure 3b). Univariate analyses identified thirty differentially abundant species after azithromycin treatment, fifteen reduced and fifteen increased (Figure 3c). The greatest reductions were found in *Lactobacillus crispatus, Klebsiella variicola, Desulfovibrio piger* and the archaeon *Methanosphaera stadtmanae*. The largest increases were in *Candidatus saccharibacteria* and *Escherichia albertii*. Additionally, seven *Acinetobacter* species were increased after treatment, including *A. baumanni, A. johnsonii, A. pitii* and *A. soli*.

**Figure 3.**
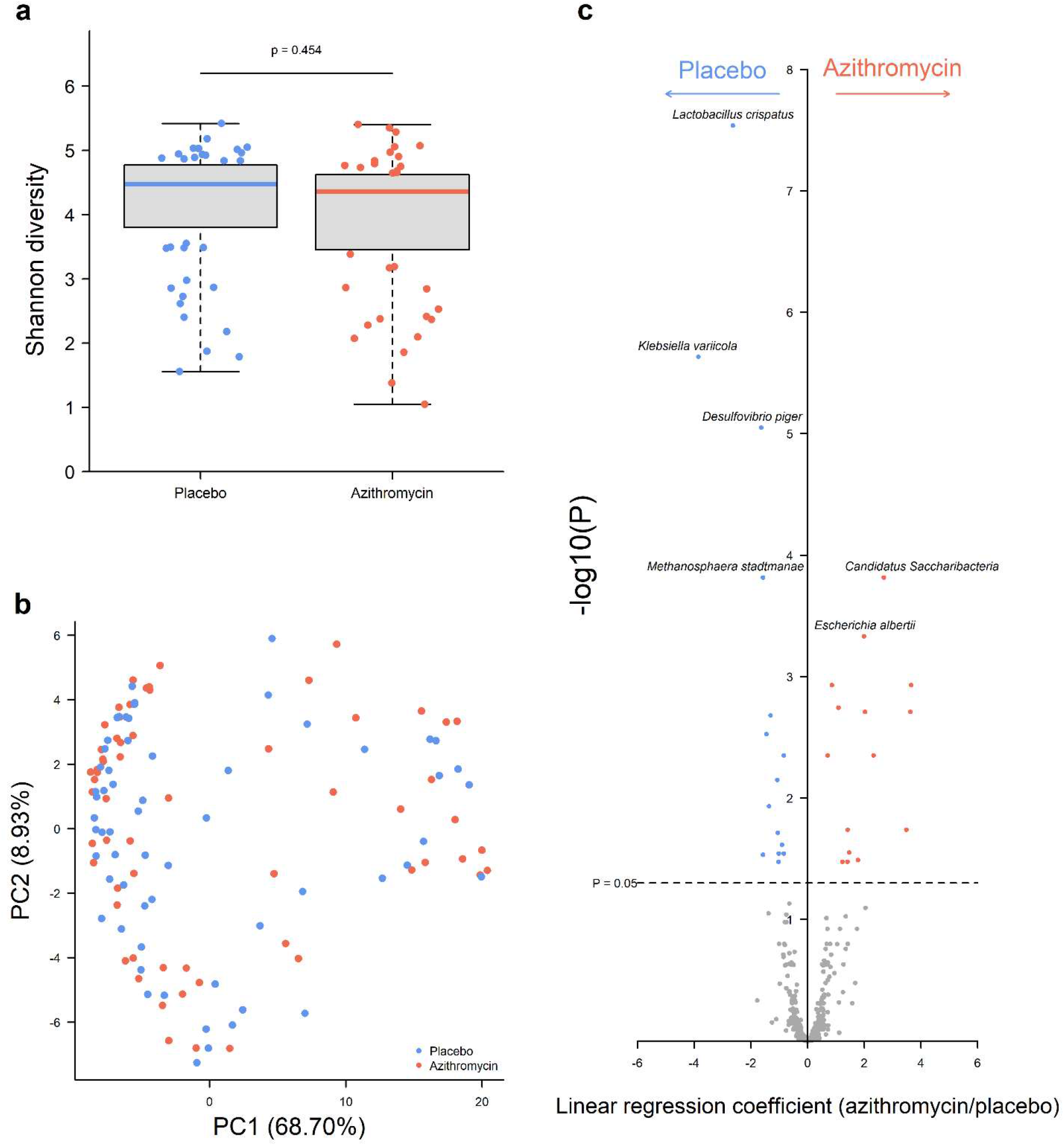
Gut microbial diversity, composition and specific bacteria in children receiving either placebo or azithromycin treatment. a. Alpha diversity, determined by Shannon’s H, at 24-month follow-up in children receiving either placebo (blue) or azithromycin (red). b. Principal component analysis (PCA) of Bray-Curtis dissimilarity between children by treatment arm. Axes labels indicate the plotted component and percentage variance explained. c. Univariate analysis of bacterial species with increased abundance (linear regression coefficient > 0) or decreased abundance (linear regression coefficient < 0) after azithromycin treatment.

We additionally examined the relationship between time since treatment and changes in the gut microbiome. Alpha diversity was not different between those treated 6-months prior or 12-24-months prior to sample collection (p = 0·114). Beta diversity showed clustering of individuals treated 6-months prior (Figure 4a). This was supported by significantly reduced abundance of thirteen bacterial species in those treated 6-months prior compared to those treated 12-24-months prior, including *Klebsiella pneumoniae* (p = 0·0002) and *Haemophilus influenzae* (p = 0·0003).

**Figure 4.**
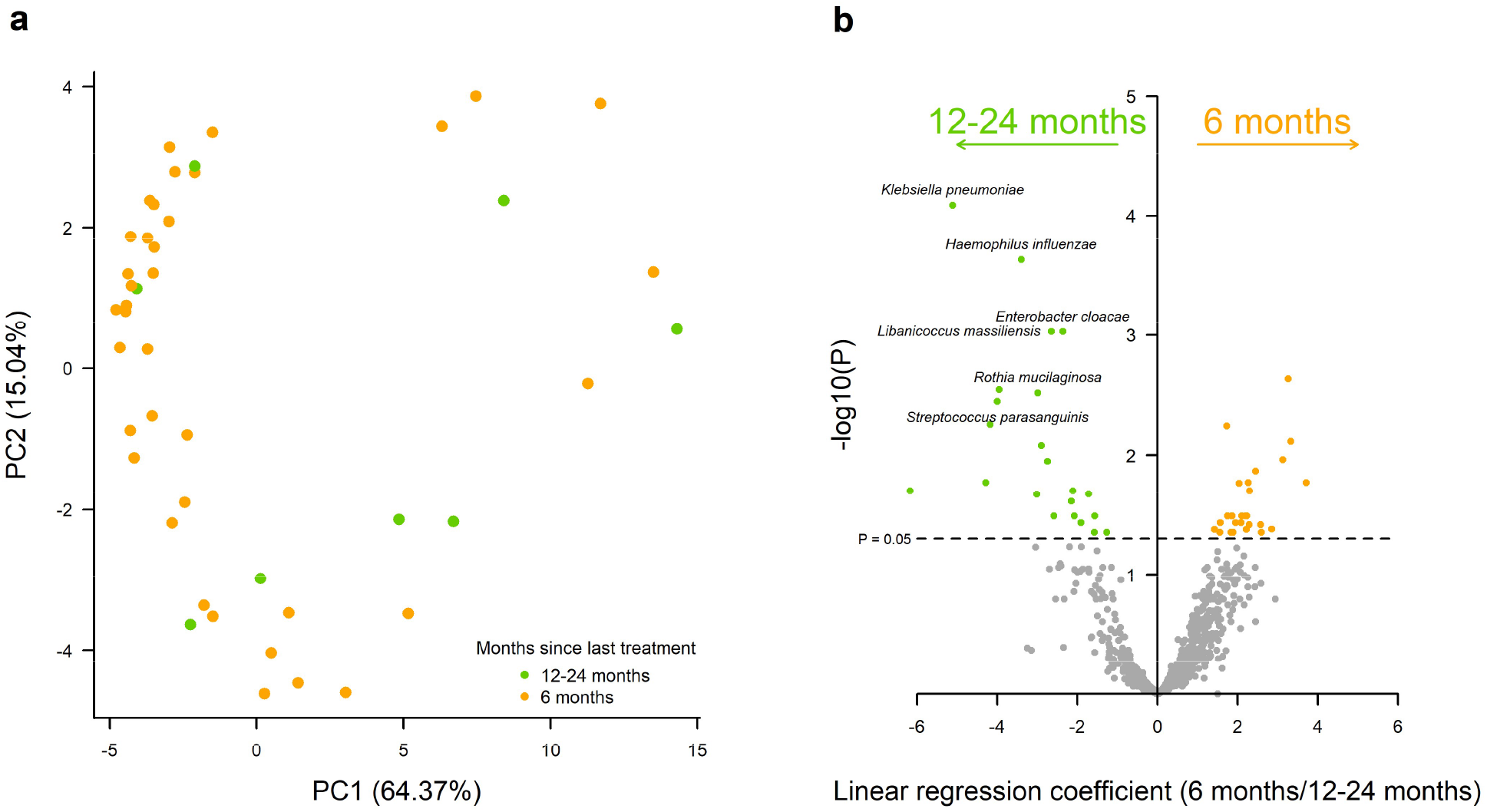
Gut microbial composition and macrolide resistance profile in children who received azithromycin 6 months or 12-24 months previously. a. Principal component analysis (PCA) of Bray-Curtis dissimilarity between children by time since last azithromycin treatment (12-24 months = green, 6 months = orange). Axes labels indicate the plotted component and percentage variance explained. b. Univariate analysis of antibiotic classes with increased evidence of resistance (linear regression coefficient > 0) or decreased evidence of resistance (linear regression coefficient < 0) in children who received azithromycin 6 months previously compared to 1-24 months previously.

To determine the impact of treatment on known causes of diarrhoea, we further investigated differences in abundance of pathogenic bacteria identified by the Global Enteric Multicenter Study (GEMS) of diarrhoeal disease in infants and young children in developing countries^38^. No genera or species was significantly different in children who received treatment (Table 3) although there was a trend was towards increased abundance in treated children for 4/5 pathogens. One of the pathogenic bacteria, *Vibrio cholerae*, was not identified in this study. Combined abundance of pathogenic bacteria was also not significantly different in children who received treatment, although the trend was again towards increased abundance in treated children.

**Table 3.**
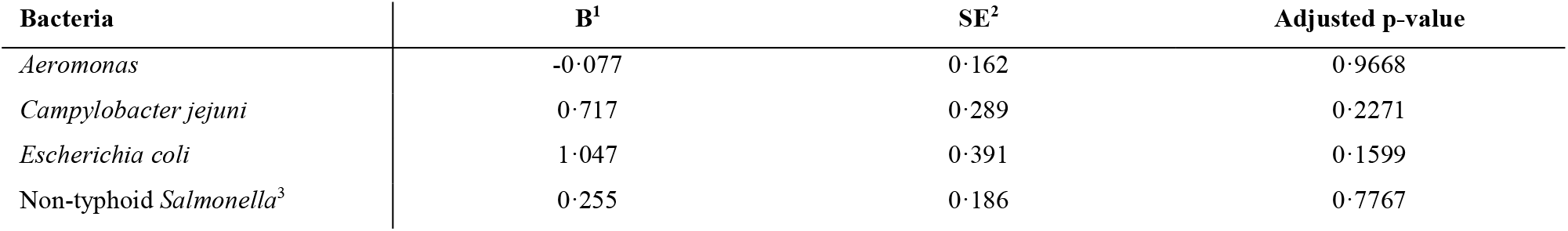
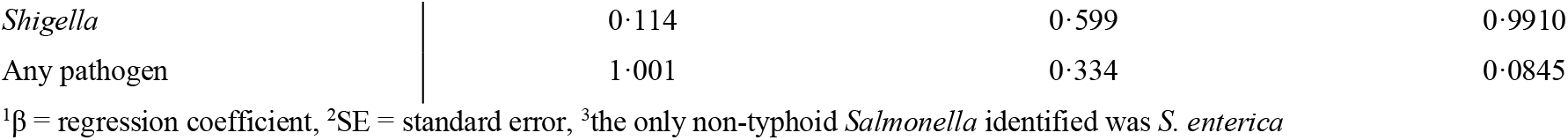
Impact of treatment on known bacterial causes of diarrhoea.

| Bacteria | B<sup>1</sup> | SE<sup>2</sup> | Adjusted p-value |
| --- | --- | --- | --- |
| *Aeromonas* | -0.077 | 0.162 | 0.9668 |
| *Campylobacter jejuni* | 0.717 | 0.289 | 0.2271 |
| *Escherichia coli* | 1.047 | 0.391 | 0.1599 |
| Non-typhoid *Salmonella*<sup>3</sup> | 0.255 | 0.186 | 0.7767 |
| *Shigella* | 0.114 | 0.599 | 0.9910 |
| Any pathogen | 1.001 | 0.334 | 0.0845 |

<sup>1</sup> $\beta$ = regression coefficient, <sup>2</sup>SE = standard error, <sup>3</sup>the only non-typhoid *Salmonella* identified was *S. enterica*

The GEMS study specifically identified enteroaggregative (EAEC), enteropathogenic (EPEC) and enterotoxigenic (ETEC) *E. coli* strains as primary causes of diarrhoea. More than 90% of *E. coli* reads here could not be classified at the strain-level; approximately 300,000 *E. coli* reads could be classified to the strain level and were analysed further. All three pathogens showed a trend towards increased abundance in treated children, significantly for EPEC (p = 0·0001, EAEC; p = 0·2970, ETEC; p = 0·8835).

## Discussion

This study utilised metagenomic sequencing to examine the impact of azithromycin treatment on carriage of macrolide-resistance bacteria in the gut. Additionally, this study investigated changes in the gut microbiome after treatment. There was significant evidence for increased macrolide resistance after treatment, the effect tended to be greater in children treated more recently. Gut microbial diversity and global community composition was not impacted by treatment, in agreement with 16S rRNA profiling of a separate longitudinal cohort of children enrolled within MORDOR in Malawi. However, individual species were differentially abundant after treatment, including the putative human enteropathogen *Escherichia albertii*.

Macrolide resistance increased after four biannual rounds of azithromycin treatment. Individuals with the highest proportion of bacteria carrying macrolide resistance had mostly been treated 6-months prior to sampling as opposed to 12-24-month prior. This effect of time since treatment was independent of number of treatments received. This suggests that impact of community-wide distribution of azithromycin on carriage of macrolide resistance may be transient. A study of children aged < five years in rural Tanzania surveyed macrolide resistance in faecal *E. coli* and found a related pattern following community-wide distribution of azithromycin for trachoma control^16^. The proportion of macrolide-resistant isolates increased sharply in the first month following MDA (16·3% to 61·2%) but had declined significantly by six months (31·3%). Studies of perturbation of the gut microbiome following azithromycin treatment consistently find detectable changes within a few days and that can last up to six months^39,40^, however, longer-lasting impact varies between studies^41,42^. Future work should ideally include sampling within days of treatment and follow-up beyond two years to determine the immediate and enduring impact of azithromycin on faecal carriage and emergence of macrolide resistance.

Prevalence of carriage of at least one macrolide resistant bacterium was higher in this study conducted in Malawian villages compared to those from Niger. At baseline in Niger, the majority of villages had no evidence of macrolide resistance, assessed by metagenomics, in either arm^26^. By contrast, 58/60 individuals (across both arms) sampled at baseline in Malawi had evidence of macrolide resistance. There is preliminary data from Tanzania suggesting availability of azithromycin at local pharmacies can impact community-level prevalence of antimicrobial resistance^43^. These Tanzanian communities had received four consecutive years of MDA and were revisited four years after cessation. At this time, there was a trend towards increased prevalence of macrolide-resistant faecal *E. coli* and nasopharyngeal *S. pneumoniae* in hamlets where azithromycin was locally available. Despite the differences in baseline prevalence of carriage of macrolide resistant bacteria, the impact of treatment was relatively consistent between the Malawi and Niger. Both reported a small reduction in gut microbial diversity after treatment and increased evidence of macrolide resistance, although the latter was determined at the village-level rather than individual-level. However, it is possible that higher baseline levels of resistance to macrolides in Malawi were a factor in the significantly reduced impact of treatment on childhood mortality compared with Niger. These hypothesis could be further evaluated by determining pre-treatment prevalence of macrolide resistance in the Tanzanian communities enrolled in MORDOR, where the lowest impact of treatment was reported^21^ and assessment of antibiotic availability in rural Malawi.

In agreement with findings from Niger, we detected no significant changes in prevalence of resistance to non-macrolide antibiotics after four rounds of treatment^26^. However, after a further two rounds of treatment in Nigerien communities^27^, resistance to several classes of non-macrolide antibiotics was increased, this was maintained after two additional treatment rounds for β-lactams. Prevalence of resistance to aminoglycosides and trimethoprim was significantly greater after six rounds of treatment in Niger; resistance to these two antibiotic classes was non-significantly increased after four rounds of treatment in our study. The most common explanation for this effect is multi-drug resistant bacteria, driven by shared mechanisms of resistance or genetic linkage^44^, however, carriage of aminoglycoside and trimethoprim resistance was independent of macrolide resistance in this study. It is possible that increases in off-target antibiotic resistance in the gut microbiome of children from studied Malawian communities would reach significance with further rounds of treatment, as reported in Niger.

Five of six bacteria highlighted as common causes of diarrheal disease by the Global Enteric Multicenter Study (GEMS) of infants and young children in developing countries were detectable in this study^38^. While none were significantly impacted by treatment, combined abundance of these diarrhoeal pathogens was higher after treatment, although this did not reach significance. Focusing on pathogenic *E. coli* strains demonstrated a significant increase in abundance of enteropathogenic *E. coli* (EPEC) after treatment. Strain-level results should be treated with caution as the majority of reads were not classifiable beyond species-level. A similar increase was observed in *Escherichia albertii*, a close relative of *E. coli*^45^. Recent data suggests many gastrointestinal infections classified as *E. coli* infections may be the related *E. albertii*^46,47^. *E. albertii* possesses many of the virulence factors found in EPEC and multi-drug resistant isolates have been recovered from clinical samples^48,49^. Importantly, there was no evidence of macrolide resistance in EPEC or *E. albertii* sequences at 24-month follow-up in either arm of the study; it is likely that abundance of this pathogen increased as an effect of treatment rather than it being resistant. For example, it is possible that higher azithromycin susceptibility of other pathogens such as *Campylobacter, Salmonella* and *Shigella* may indirectly lead to increased abundance of EPEC and *E. albertii*^50^. Targeted higher-resolution sequencing of *Escherichia* to accurately differentiate species and identify strains of *E. coli* would be of value.

In this study, the abundance of several *Acinetobacter* species increased after treatment, which can be partly explained by their intrinsic resistance to macrolides^51^. Despite this, the prevalence of the opportunistic pathogens *A. baumanni, A. johnsonii, A. pitii* and *A. soli* is concerning. *A. baumanni* infections are often hospital-acquired with the majority of community-acquired infections presenting in individuals with underlying health conditions^52,53^. High mortality rates and rapid emergence of antimicrobial resistance suggest this species requires consideration as a serious human pathogen. This was highlighted in this study by evidence of two macrolide-efflux genes (*mphE* and *msrE*) in an *A. baumanni* isolate from a child in the azithromycin treatment arm. Further studies are required to elucidate the impact of these species and whether gut residence is evidence of an active infection or a reservoir for infections at other sites.

The findings presented here are limited to a single geographical zone in the Mangochi district of Malawi, which may limit extrapolation to wider populations. Further to this, participation in faecal sampling was incomplete as approximately 70% of enrolled children provided samples. Given the observed individual-level heterogeneity in this study, it is possible more complete sampling of enrolled children would have led to different outcomes. However, the overlap with results from Niger suggest these effects may be consistent across study sites. Additionally, very few of the individuals sampled herein were aged 1-5 months, the group which saw the greatest reduction in mortality after treatment. It is possible that treatment may have a different impact on the gut microbiome in these younger children.

In summary, this study found significant changes in the antimicrobial resistance profile and gut microbiome after four biannual rounds of azithromycin. Carriage of macrolide resistance was increased by treatment, this effect was more pronounced in recently treated children. Abundance of the putative human enteropathogen *E. albertii* was increased after treatment, as well as several opportunistic *Acinetobacter* pathogens. Given that multiple treatments enhanced the beneficial effects of azithromycin treatment and more recent treatment favoured increases in macrolide resistance, consideration should be given to the number of treatments and administration schedule to ensure benefits continue to outweigh costs in antimicrobial resistance.

## Data Availability

All sequence data are available from the European Bioinformatics Institute (EBI) short read archive (PRJEB42363).

## Contributors

JDH, SB, KM, KK, RLB and MJH contributed to study design. JDH, SB, KM, KK and RLB contributed to data collection. HP, SB, RS and MJH contributed to data analysis. All authors interpreted the findings, contributed to writing the manuscript, and approved the final version for publication.

## Acknowledgements

We would like to acknowledge the contributions of field staff at Blantyre Institute of Community Outreach and laboratory staff at College of Medicine, Blantyre, Malawi.

